# Levels of high-sensitive troponin T and mid-regional pro-adrenomedullin after COVID-19 vaccination in vulnerable groups: a prospective study on subtle and persistent cardiovascular involvement

**DOI:** 10.1101/2024.05.10.24307207

**Authors:** Martin Möckel, Samipa Pudasaini, Ngoc Han Le, Dörte Huscher, Fabian Holert, David Hillus, Pinkus Tober-Lau, Florian Kurth, Leif Erik Sander

## Abstract

**Background:** This study examines potential, subtle and persistent adverse effects of COVID-19 vaccines on the cardiovascular system. Vaccine-associated myocardial injury was analysed by measuring high-sensitive troponin T (hsTnT); mid-regional pro-adrenomedullin (MR-proADM) levels were evaluated to assess endothelial dysfunction.

**Methods:** This was a prospective study with a vulnerable population of healthcare workers (HCWs) and elderly patients (> 70 years) who were vaccinated with either one dose of ChAdOx1 nCov-19 adenoviral vector vaccine (AZ) followed by one dose of the BNT162b2 messenger RNA vaccine (BNT), or with two doses of BNT (12^th^ of January - 30^th^ of November 2021). HsTnT and MR-proADM were measured in blood samples at three visits (V_1_: 1^st^ immediately before vaccination; V_2, 3_: 3-4 weeks after 1^st^ and 2^nd^ vaccination). HsTnT of HCWs was compared to a healthy reference population.

**Results:** N=162 volunteers were included (V_1_=161; V_2_, V_3_=162 each). N=74 (45.7%) received AZ/BNT and n=88 (54.3%) received BNT/BNT (elderly: n=20 (12.3%), HCWs: n=68 (42.0%)). Median hsTnT levels were 4ng/L, 5ng/L and 4ng/L (V_1_-V_3_) for AZ/BNT and at 5ng/L, 6ng/L and 6ng/L (V_1_-V_3_) for BNT/BNT. Compared to the reference population (n=300), hsTnT was significantly higher at all visits for both vaccination groups (p<0.01), without differences between the AZ/BNT and BNT/BNT cohort. MR-proADM values were 0.43nmol/L, 0.45nmol/L, 0.44nmol/L (V_1_-V_3_) in the AZ/BNT cohort and 0.49nmol/L, 0.44nmol/L, 0.47nmol/L for BNT/BNT, respectively. Change of median hsTnT and MR-proADM between visits did not show significant increases. One HCW case had a permanent and three a transient hsTnT increase ≥14ng/L.

**Conclusion:** With one individual exception, no overall subtle, persistent cardiovascular involvement was observed after the 2^nd^ COVID-19 vaccination.

**Structured graphical abstract:** Summary of the vaccination scheme and visiting points in the study population (of HCWs and seniors > 70 years) between the 12^th^ of January and the 30^th^ of November 2021. The results showed no overall subtle, chronic myocardial or vascular involvement in our COVID-19 vaccinated cohorts. <u>Abbreviations:</u> AZ ChAdOx1 nCov-19 adenoviral vector vaccine from Astra Zeneca, BNT BNT162b2 messenger ribonucleic acid vaccine from BioNTech, EDTA Ethylenediaminetetraacetic acid, HCWs health care workers, hsTnT high-sensitive troponin T, mid-regional pro-adrenomedullin, V_1_-V_3_ visiting times 1-3, w week(s).

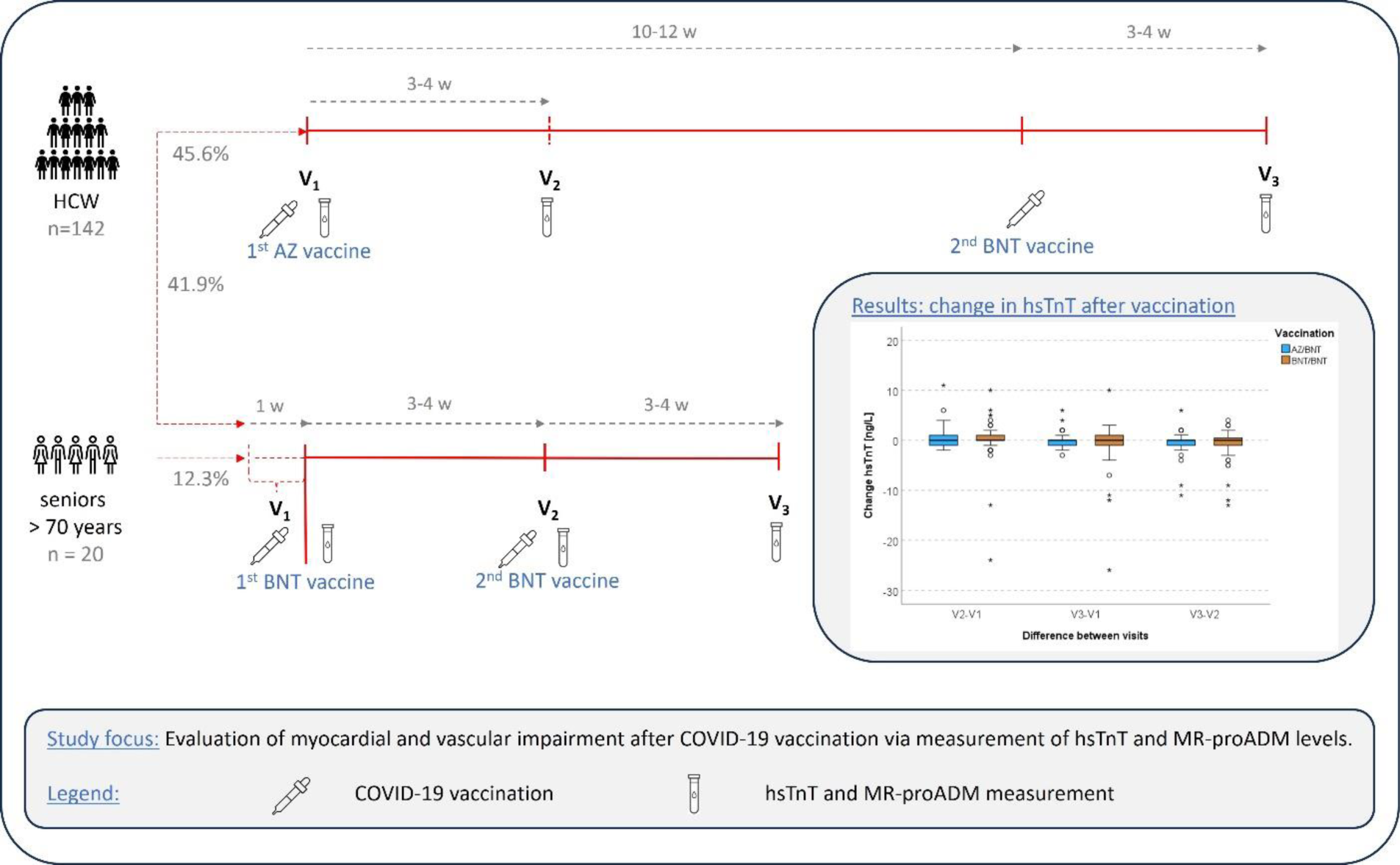

## Introduction

Coronavirus disease 2019 (COVID-19) vaccines are generally safe and effective in reducing the severity of SARS-CoV-2 infections and their complications (1, 2). About 70.6% of the world’s population has received at least one vaccine dose, and over 13 billion doses have been given worldwide (3). To ensure the safety of COVID-19 vaccines, short- and long-term adverse events are continuously monitored by active and passive pharmacovigilance and post-marketing studies and vaccine safety is intensely studied (4, 5). The International Coalition of Medicines Regulatory Authorities, representing 38 regulatory agencies, recently issued a joint statement on reassuring safety data showing that COVID-19 vaccines have a very good safety profile in all age groups, including children and people with underlying medical conditions (6). Rare adverse events include complications of the cardiac system, like COVID-19 vaccine-associated myocarditis and pericarditis, as well as adverse effects on the vasculature, such as an impaired endothelial function as well as arterial and venous thrombosis (7–10).

### COVID-19 vaccination and the cardiac system

Although rare, myo- and pericarditis are the most frequent adverse cardiac events following COVID-19 vaccinations (11, 12) and were first reported in April 2021 (12). Today, incidence of myo- and pericarditis is (region-specifically) estimated at 1 to 11 per 100,000 vaccinees (12, 13). Incidence is highest among young adult males, particularly following the second dose of Moderna (MOD) or BioNTech/Pfizer (BNT) messenger ribonucleic acid (mRNA) vaccine (12). In patients with mRNA-vaccine related myo- and/or pericarditis, cardiomyocyte necrosis was almost always detectable, as determined by measurements of the cardiac-specific biomarker high-sensitive troponin T (hsTnT) (14, 15). For instance, in a study by Oster *et al.*, elevated hsTnT was observed in 97.9% (792/809) of COVID-19 vaccine-associated cases of myo- and/or pericarditis, as well as further major adverse cardiac events (14). Hence, hsTnT measurement is of high relevance for the diagnostic identification of patients with vaccine-related cardiac adverse events.

So far, cardiomyocyte injury in COVID-19 vaccine recipients was primarily analysed in study populations that presented with clinically manifest and acute cardiac symptoms (7). Yet, it has rarely been investigated if and to what extent a subtle, chronic post-vaccine myocardial involvement occurs in a vaccinated population even in the absence of typical symptoms. Mainly three studies actively monitored of hsTnT levels in a BNT-vaccinated cohort. Mansanguan *et al.* analysed 301 adolescents of whom four had signs of a subclinical myocarditis, one suffered a clinically manifest myopericarditis and two a pericarditis after the 2^nd^ BNT162b2 vaccine. Here, cardiac biomarkers were measured at baseline, day 3, 7 and 14 (optionally) (16). Levi and colleagues observed vaccine-associated cardiomyocyte injury in two participants (0.6%), with one being symptomatic and one asymptomatic, 2 to 4 days after having received their 4^th^ BNT dose (17). Lastly, Buergin *et at.* detected the highest rate of 2.8% (women: 20/777; men: 2/777) in their hospital employee cohort presenting with a vaccine-induced myocardial injury on day 3 after receipt of MOD mRNA booster (18). All three studies described cardiac involvement as mild and transient and mainly defined it as an acute hsTnT elevation above the 99^th^ percentile of upper reference limit (URL) (16, 18, 19). Electrocardiograms and echocardiography were normal in the majority of affected participants (17, 18).

In order to further strengthen safety data and confidence in COVID-19 vaccines, additional research is warranted to assess possible subtle and subclinical myocardial damage after COVID-19 vaccination, also with regards to varying vaccination regiments (vector versus mRNA or heterologous vaccination). Furthermore, as emphasised in a recent editorial by Levi *et al.*, longitudinal observations of hsTnT levels over time, e.g. including pre-vaccination status and follow-up samples after booster vaccination, would be of interest to evaluate if any occurring subclinical cardiac injury is likely self-limiting or becomes chronic (17).

### COVID-19 vaccination and the vascular system

Mainly one study has investigated possible acute effects of COVID-19 vaccines on the vascular system (20). The authors observed a short-term deterioration of endothelial function in the first 24 hours after the 2^nd^ BN vaccine dose. The extent of endothelial involvement was described as far lower than in COVID-19, which in turn has been linked to endothelial cell infection and endotheliitis (20). In COVID-19 patients, the biomarker mid-regional pro-adrenomedullin (MR-proADM), an endothelium-related peptide with vasodilatory properties, was used to evaluate the severity of endothelial dysfunction (21). Measurements of MR-proADM levels to assess possible short- and long-term endothelial injury in COVID-19 vaccine recipients have not been analysed so far.

### Study focus

By conducting a prospective observational study, we investigated subacute, subtle, and subclinical cardiomyocyte injury in two risk populations with follow-ups until 4 weeks (optionally longer) after completion of the original COVID-19 basic vaccination regimen. We hypothesize that hsTnT levels remain unchanged during follow-up, indicating no significant rise in myocardial injury in the short- and long-term compared to baseline. Secondly, we sought to analyse endothelial function based on levels of MR-proADM throughout the study period. With these two approaches, we aimed to address current knowledge gaps concerning chronic cardiovascular effects of COVID-19 vaccination.

## Methods

### Study design and study population

The participants were recruited within the multicentre prospective observational studies, EICOV, COVIMMUNIZE and COVIM and consisted of health care workers (HCWs) of Charité – Universitätsmedizin Berlin as well as elderly/senior patients (> 70 years) at a general practitioner’s office in Berlin, Germany; two different types of risk populations in the pandemic. While vulnerability of HCWs is caused by their regular exposition to infectious patients, elderly mainly fall into this category due to their compromised immune response and multi-morbidity (22, 23). All participants received their first two COVID-19 vaccinations between 12^th^ of January and 22^nd^ of June 2021 with either a combination of AZ (AstraZeneca)/BNT (only for HCWs) or BNT/BNT (HCWs or elderly). Booster vaccinations (3^rd^ and 4^th^) were only mRNA-based (BNT or MOD). Key inclusion criteria were the ability to give written informed consent by the participants or via their legal representative, no contraindications to receiving a COVID-19 vaccination and an age of ≥18 years at the time of enrolment.

Three main visit timepoints (V_1_-V_3_) were conducted. V_1_: at baseline, i.e., 7 to 0 days before the 1^st^ vaccination; V_2_: 3-4 weeks after the 1^st^ vaccination and V_3_: 3-4 weeks after the 2^nd^ vaccination (Figure (Fig.) 1). Prior work on this topic has shown that cardiovascular biomarkers, if elevated in the acute setting, usually rise and fall within 1-2 weeks after vaccinations. By choosing an interval of 3-4 weeks post-vaccination for biomarker measurement, we aimed to assess subacute or persistent hsTnT elevation (16). Participants that opted to participate in the follow-up study COVIM-Boost, a 4^th^ visit (V_4_) and 5^th^ visit (V_5_) was captured and is reported here for HCWs as part of an ancillary analysis up to 50 days after the 3^rd^ and 4^th^ vaccination (with either BNT or MOD), respectively (Supplementary Fig. 1). At all visits, serum and ethylenediaminetetraacetic acid (EDTA) blood samples were collected. MR-proADM was measured in EDTA plasma [nmol/L] on the Kryptor Compact Plus device (Thermo Fisher Scientific, Waltham, MA, USA). The clinically relevant cut-off was set at ≥ 0.75 nmol/L (24). Depending on the sample availability, EDTA or serum samples were used for measuring hsTnT (via Cobas e801, Roche Diagnostics, Mannheim, Germany). In the diagnostic exclusion procedure of a myocardial infarction, relevant hsTnT levels are defined as being > the 99^th^ percentile of URL (25). This definition was also adopted by prior studies with a research question similar to ours (16–18). However, in a study by Barbier *et al.* the authors pointed out that new, subclinical cardiomyocyte injury, detected in late enhancement magnetic resonance imaging, may only be reflected in minor troponin elevations (of 4.1ng/L to 5.9ng/L) and slight increases also correlate with a negative prognosis (26). Additionally, in cases of SARS-CoV-2 infections with cardiac involvement, troponinemia was likewise mostly mild (27). Therefore, to also capture a possibly subtle but relevant troponin increase, we focused on all changes in hsTnT levels in the course of the visits and put it in relation to the initial baseline hsTnT value of the respective participant. Cases with a hsTnT elevation > the 99^th^ percentile cut-off value were presented and interpreted separately. HsTnT levels below the limit of detection (3ng/L), were set to 2.9ng/L for analyses.

**Figure 1.**
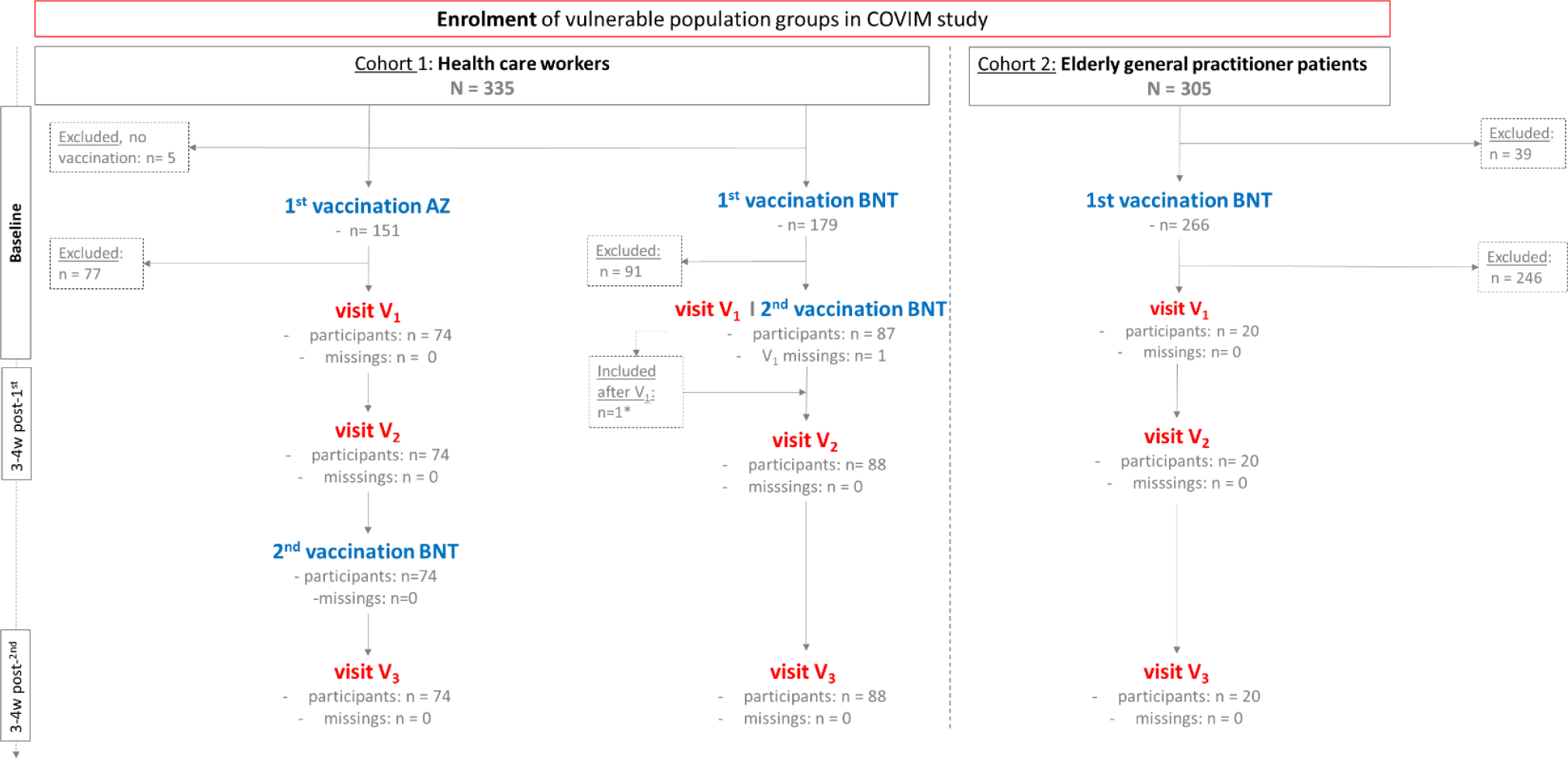
Study population diagram. Recruitment of vulnerable study population with cohort 1 including health care workers and cohort 2 including elderly general practitioner patients. Vaccinations with either a heterogenous combination of AZ/BNT or a homologous scheme of BNT/BNT were applied and three main visits V_1_ to V_3_ were performed. * One hospital employee of the BNT/BNT cohort did not participate in the enrolment visit V_1_ but received both BNT vaccinations regularly and took part in the visits V_2_, V_3_ and is, therefore, included in this study. <u>Abbreviations</u>: AZ ChAdOx1 nCov-19 adenoviral vector vaccine from Astra Zeneca, BNT BNT162b2 messenger ribonucleic acid vaccine from BioNTech, V_1_-V_5_ visiting times 1-5, w week(s).

Demographic and clinical characteristics of all participants, including age and sex, comorbidities and pre-medication were collected at enrolment and used for comparison with the reference cohort.

### Reference population

As a reference population, participants of the BIC-1 (**B**iomarkers **i**n **C**ardiology **1**) study were used. This included HCWs of Charité – Universitätsmedizin Berlin that were recruited between 6^th^ of July 2007 and 5^th^ November 2007. Clinical exclusion criteria were the presence of a heart failure, other heart diseases, kidney or metabolic diseases and the intake of permanent medication. Therefore, this cohort was defined as a presumably healthy reference and was used for comparison of hsTnT values with our HCWs in the vaccination groups. Further inclusion criteria include a full employment contract, age of 18 years or older and no direct dependence on the study leader were required for inclusion. In this cohort, one visit (V_R-1_) was performed. HsTnT was measured in lithium-heparin blood samples on the same analyser module as in our vaccination cohorts (Cobas e801, Roche Diagnostics, Mannheim, Germany).

### Hypothesis

The primary hypothesis was that the distribution of hsTnT remains unchanged across all vaccination groups at the visits V_1_ to V_3_. Therefore, we assumed that neither the combination of AZ/BNT, nor the BNT/BNT scheme would lead to a significant elevation of hsTnT compared to baseline levels at enrolment visit V_1_. Secondly, we expected no relevant chronic endothelial dysfunction to be detected in the sense of a significant median difference between V_1_ to V_3_ of MRpro-ADM values in the observed time span.

### Statistical analysis

A basic description of the vaccination and reference groups was performed, including characteristics of sex, age, Body-Mass-Index (BMI), smoking status, comorbidities, and pre-medication at enrolment. If not indicated otherwise, metric values are presented as median with the associated interquartile range (IQR). Due to the skewed distribution of hsTnT and MR-proADM values, non-parametric tests were performed. The Mann-Whitney U test was applied to test the difference of hsTnT and MR-proADM levels between the vaccination groups, and the Wilcoxon signed rank test to investigate individual change between the visits for both biomarkers (differences V_2_-V_1_, V_3_-V_1_, V_3_-V_1_). Correlation analyses were performed separately for sex between hsTnT and MR-proADM at each visit, and for both hsTnT and MR-proADM with age at V_1_. Spearman’s correlation coefficients were reported. Comparisons of hsTnT values between the reference group and the two vaccination cohorts (solely HCWs included here) were performed with the Kruskal-Wallis-test and pairwise post-hoc tests with Bonferroni correction. Mixed-effects models were calculated to examine effects of age (per 10 years), sex, and the type of COVID-19 vaccination on the course of hsTnT and MR-proADM values, with random effects for the intercept. Since most visits were performed within a very close time window, the course of hsTnT and MR-proADM values were further explored in mixed-effects regression models treating the visits as repeated measurements, again including random effects for the intercept. In addition, mixed-effects regression models for propensity score-matched groups were calculated to examine effects of the vaccinations while controlling for potential confounders.

All analyses were performed using IBM SPSS Version 29 (IBM Corporation, Armonk, NY, USA) and R version 4.3.1 (2023-06-16 ucrt). A p-value <0.05 was considered significant; for pairwise subgroup comparisons or parallel tests in visits V_1_ to V_3_, p-values were adjusted for multiple testing.

### Ethical approval and study registration

The COVIM study (EA4/245/20), its preceding studies EICOV (EA4/245/20) and COVIMMUNIZE (EA4/244/20), the follow-up study COVIM-Boost (EA4/261/21) and the BIC-1 study (EA2/030/07) were approved by the ethics committee of Charité – Universitätsmedizin Berlin. BIC-1 was registered in the German register for clinical studies (DRKS-ID DRKS00000310) and COVIM in the European clinical trial register (EudraCT-2021–001512–28).

## Results

### Characteristics of the study population

Information on basic characteristics is provided in Table (Tab.) 1. In total, the study population consisted of 162 people in the vaccination group, including 142 (87.7%) HCWs and 20 (12.3%) elderly patients (solely in the BNT/BNT group). Except for one person who missed the 1^st^ visit, all participants were seen (at least) three times. Altogether, 74 (45.7%) participants received the combination of AZ/BNT, while 88 (54.3%) participants were vaccinated according to regular BNT/BNT regimen. In the AZ/BNT group, 41 (55.4%) participants and in the BNT/BNT population 49 (55.7%) were female, respectively. The median age was 33.2 years in the AZ/BNT and 37.6 years in the BNT/BNT group. Median BMI was 23.9 kg/m^2^ and 23.8 kg/m^2^, respectively. Out of 148 available answers, 16 reported being either former current smokers.

With regards to pre-medication, 9 (5.6%, 7/9 from the BNT/BNT cohort) reported receiving immunosuppressive therapy and 4 (2.5%, 3/4 from the BNT/BNT group) participants reported taking corticosteroids at enrolment. Furthermore, 14 (8.6%) participants reported taking antihypertensive drugs, 3 (1.9%) anticoagulants and 4 (2.5%) antiplatelet drugs as a permanent medication, the majority of participants on this medication belonging to the elderly group (see Tab. 1.)

**Table 1.** Baseline characteristics of the study cohort and reference group.

| Characteristics |  | COVIM study |  |  |  | BIC-1 study |
| --- | --- | --- | --- | --- | --- | --- |
|  | All participants | AZ/BNT | BNT/BNT |  |  | Reference group |
|  |  |  | In total | HCWs | elderly |  |
| Numbers (n) | 162 | 74 | 88 | 68 | 20 | 300 |
| Patient characteristics |  |  |  |  |  |  |
| Age, years<sup>#</sup> | 35.6 (30.0-54.1) | 33.2 (29.7-45.3) | 37.6 (31.1-59.6) | 34.2 (29.7-44.0) | 83.7 (80.7-87.0) | 39.0 (33.0-46.0) |
| Sex, female | 90 (56%) | 41 (55%) | 49 (56%) | 34 (50%) | 15 (75%) | 150 (50%) |
| BMI, kg/m<sup>2</sup> <sup>#1</sup> | 23.9 (21.5-25.8) | 23.9 (21.7-25.7) | 23.8 (21.3-26.1) | 24.1 (21.6-26.1) | 23.2 (21.5-26.0) | 24.9 (22.4-27.7) |
| Former smokers | 6/148 (4.0%) | 2/72 (2.7%) | 4/76 (5.2%) | 4/56 (7.14%) | 0/20 (0%) | 61/300 (20.3%) |
| Current smokers | 10/148 (6.7%) | 4/72 (5.5%) | 6/76 (7.8%) | 5/56 (8.9%) | 1/20 (5%) | 105/300 (35.0%) |
| Comorbidities* |  |  |  |  |  |  |
| Cardiovascular diseases | 21/162 (13.0%) | 4/74 (5.4%) | 17/88 (19.3%) | 3/68 (4.4%) | 14/20 (70%) | n. a. |
| Hypertonus | 20/162 (12.3%) | 4/74 (5.4%) | 16/88 (18.2%) | 3/68 (4.4%) | 13/20 (65%) |  |
| Heart insufficiency | 4/162 (2.5%) | 0 | 4/88 (4.5%) | 0 | 4/20 (20%) |  |
| Heart rhythm disorder | 3/162 (1.9%) | 0 | 3/88 (3.4%) | 0 | 3/20 (15%) |  |
| Myocardial infarction, Angina pectoris, peripheral artery disease, carotid stenosis | 3/162 (1.95%) | 0 | 3/88 (3.4%) | 0 | 3/20 (15%) |  |
| Chronic lung diseases | 10/162 (6.2%) | 4/74 (5.4%) | 6/88 (6.8%) | 4/68 (5.9%) | 2/20 (10%) |  |
| Asthma | 6/162 (3.7%) | 3/74 (4.1%) | 3/88 (3.4%) | 3/68 (4.4%) | 0 |  |
| Chronic Bronchitis | 1/162 (0.6%) | 0 | 1/88 (1.1%) | 1/68 (1.5%) | 0 |  |
| COPD | 3/162 (1.9%) | 1/74 (1.4%) | 2/88 (2.3%) | 0 | 2/20 (10%) |  |
| Kidney diseases | 7/162 (4.3%) | 1/74 (1.4%) | 6/88 (6.8) | 0 | 6/20 (30%) |  |
| Cancer | 7/162 (4.3%) | 3/74 (4.1%) | 4/88 (4.5%) | 1/68 (1.5%) | 3/20 (15%) |  |
| Pre-medication* |  |  |  |  |  |  |
| Immunosuppression | 9/162 (5.6%) | 2/74 (2.7%) | 7/88 (8.0%) | 3/68 (4.4%) | 4/20 (20%) | n. a. |
| Corticosteroids | 4/162 (2.5%) | 1/74 (1.4%) | 3/88 (3.4%) | 0 | 3/20 (15%) |  |
| Antihypertensive drugs | 14/162 (8.6%) | 2/74 (2.7%) | 12/88 (13.6%) | 1/68 (1.5%) | 11/20 (55%) |  |
| Anticoagulants | 3/162 (1.9%) | 0 | 3/88 (3.4%) | 0 | 3/20 (15%) |  |
| Antiplatelet drugs | 4/162 (2.5%) | 0 | 4/88 (4.5%) | 0 | 4/20 (20%) |  |
| Baseline biomarker values |  |  |  |  |  |  |
| hsTnT (V<sub>1</sub>, V<sub>R-1</sub>), ng/dl<sup>#</sup> | 5 (4-7) | 4 (4-6) | 5 (4-8) | 5 (4-6) | 18 (11-29) | 3.6 (3.0-4.9) |
| MR-proADM (V<sub>1</sub>), nmol/L<sup>#</sup> | 0.44 (0.40-0.55) | 0.43 (0.39-0.47) | 0.49 (0.42-0.63) | 0.44 (0.40-0.54) | 0.95 (0.65-1.01) | n. a. |

\*: more than one choice possible. #: median with interquartile range (IQR). N.a.: not applicable. <sup>1</sup> BMI
values were available 147 participants in total with 73 AZ/BNT individuals and 74 BNT/BNT people.
Abbreviations: AZ ChAdOx1 nCov-19 adenoviral vector vaccine from Astra Zeneca, BMI Body-Mass-Index, BNT BNT162b2 messenger ribonucleic acid vaccine from BioNTech, COPD Chronic Obstructive Pulmonary Disease, hsTnT high-sensitive troponin T, MR-proADM mid-regional pro-adrenomedullin.

Four (5.4%) participants in the AZ/BNT cohort, and 17 (19.3%) participants in the BNT/BNT cohort reported a cardiovascular disease at baseline, with hypertension (AZ/BNT: 4/74 (5.4%); BNT/BNT: 16/88 (18.2%)) as the most frequent condition. Chronic lung diseases were present in 10 (6.2%) participants, with 4 (5.4%) being in the AZ/BNT and 6 (6.8%) participants in the BNT/BNT cohort. Other comorbidities, with 7 cases each (4.3%), included kidney disease (6/7 BNT/BNT), rheumatologic/immunological diseases (4/7 BNT/BNT), and cancer (5/7 BNT/BNT). Diabetes, dementia, cerebrovascular or chronic haematological diseases were present in 5 (3.0%) cases, or less.

### Characteristics of the reference population

The reference population included 300 individuals with a 1:1 female to male ratio. The median age was 39.0 years. For all 300 participants, hsTnT values were available at V_R-1_ for analysis. Comorbidities and pre-medication are not reported for this cohort since both were exclusion criteria (Tab. 1).

#### HsTnT measurement

The median values of hsTnT in the AZ/BNT group were 4ng/L (IQR 4-6), 5ng/L (IQR 4-6) and 4ng/L (IQR 4-6) for V_1_, V_2_, V_3_, and for the BNT/BNT cohort 5ng/L (IQR 4-8), 6ng/L (IQR 4-9) and 6ng/L (4-8.5), respectively. Separated by HCWs and elderly (BNT/BNT), the seniors showed higher median values of 18ng/L (IQR 11-29), 15ng/L (IQR 10.5-25.5) and 13.5ng/L (IQR 10-22.5) during follow-up visits. HsTnT levels at V_1_-V_3_ in the AZ/BNT group were lower compared to the BNT/BNT cohort, at all visits, including at baseline (p_V1_=0.048, p_V2_=0.003, p_V3_<0.001). The individual progression for V_1_-V_3_ is visualised in Fig. 2A, separated by vaccination regimen and sex. Median hsTnT in the reference population was 3.6ng/L (IQR 3.0-4.9, Fig. 3). For all HCWs, median hsTnT values were higher in males at all visits, including baseline (V_1_: p=0.010; V_2_: p=0.003; V_3_: p=0.006). Changes of median hsTnT values were 0ng/L (IQR −1.0 to 1.0; V_2_-V_1_), 0ng/L (IQR −1.0 to 0.0; V_3_-V_2_) and 0ng/L (IQR −1.0 to 0.0; V_3_-V_1_) in the AZ/BNT cohort (Fig. 2B). In the BNT/BNT population, changes were 0ng/L (IQR 0.0 to 1.0; V_2_-V_1_), 0ng/L (IQR −1.0 to 1.0; V_3_-V_2_) and 0ng/L (IQR −1.0 to 1.0; V_3_-V_1_). There were no individual differences of hsTnT levels between all visits (AZ/BNT: V_2_-V_1_ p=1.0; V_3_-V_2_ p=0.25; V_3_-V_1_ p=0.42; BNT/BNT: V_2_-V_1_ p=0.24; V_3_-V_2_ p=0.25; V_3_-V_1_ p=1.0). Median changes of hsTnT for HCWs and elderly of the BNT/BNT cohort separately showed significant differences between V_2_ and V_1_ for HCWs (median 0 ng/L (IQR 0.0 to 1.0), p=0.029) and between V_3_ and the two prior visits for elderly (V_3_-V_2_: median −1 ng/L (IQR −4.0 to 0.0), p=0.038; V_3_-V_1_: median −2ng/L (IQR −4.0 to 0.0), p=0.008) were visible (Fig. 2C).

**Figure 2.**
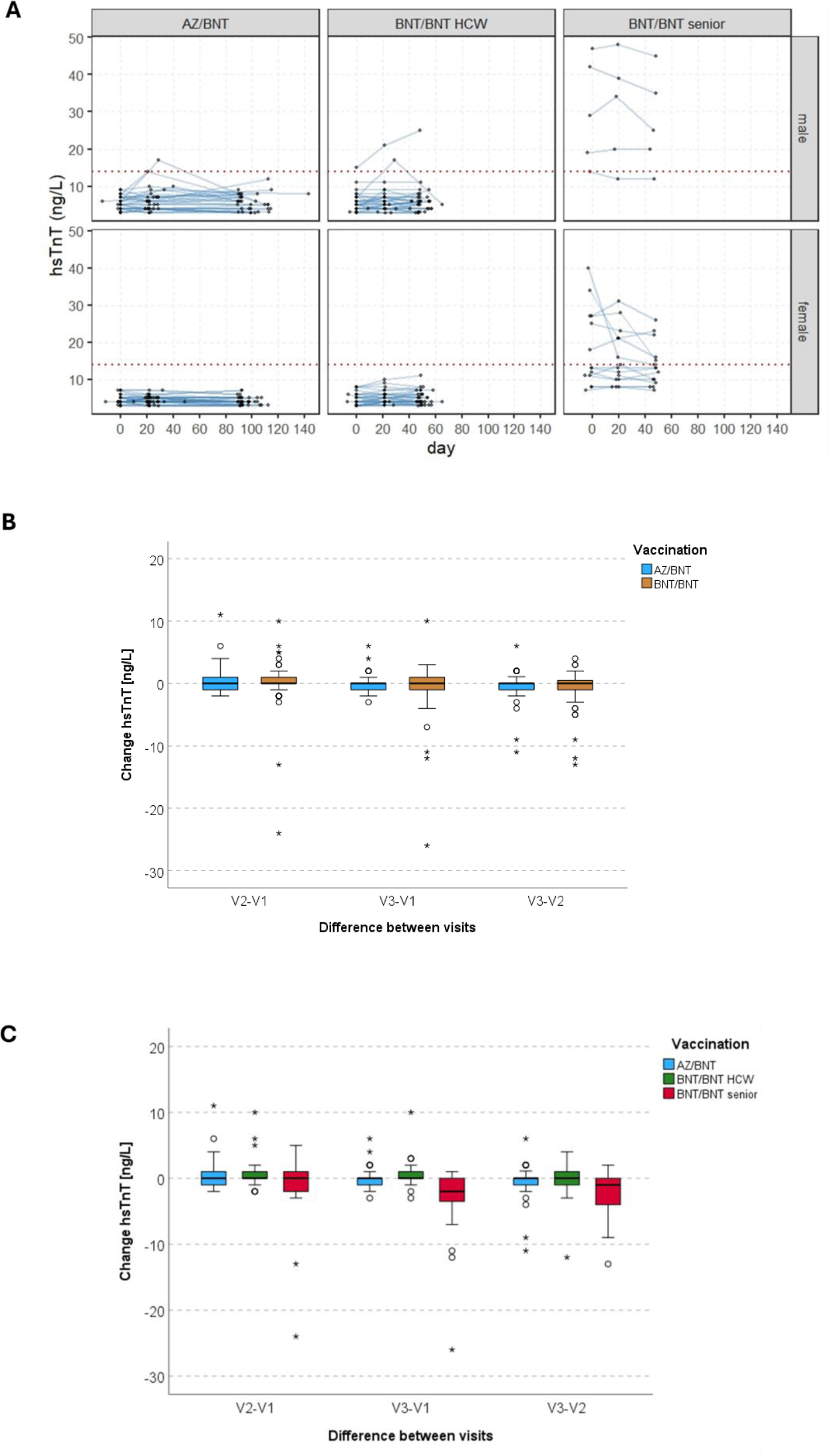
**A**. HsTnT values at the three main visiting points for participants of the AZ/BNT and BNT/BNT group (HCWs and elderly), separated by sex. The red dashed line indicates the 14 ng/L threshold. **B.** Box plots of the change of hsTnT values between each visiting time (V_1_-V_2_, V_1_-V_3_, V_1_-V_3_), separated by AZ/BNT and BNT/BNT group. **C.** Box plots of the change of hsTnT values between each visiting time (V_1_-V_2_, V_1_-V_3_, V_1_-V_3_), separated by AZ/BNT, HCW BNT/BNT and senior BNT/BNT group. <u>Abbreviations</u>: AZ ChAdOx1 nCov-19 adenoviral vector vaccine from Astra Zeneca, BNT BNT162b2 messenger ribonucleic acid vaccine from BioNTech, hsTnT high-sensitive troponin T, V_1_-V_3_ visiting times 1-3.

**Figure 3.**
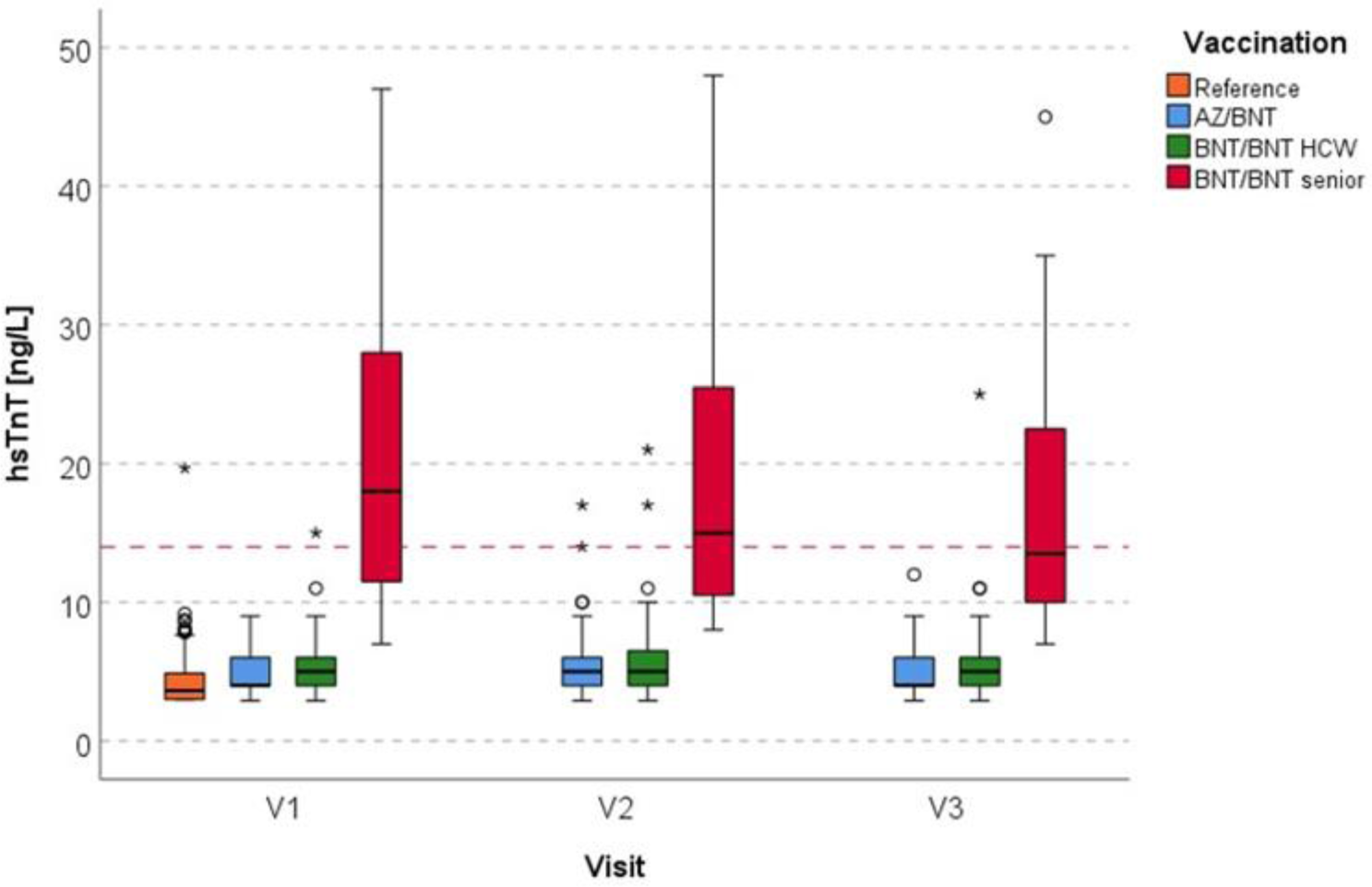
Box plots of hsTnT levels of the reference group (orange) compared to the AZ/BNT group (blue) and the BNT/BNT group (HCWs: green; elderly: red) at V_1_ to V_3_. The red dashed line indicates the 14 ng/L threshold. <u>Abbreviations</u>: AZ ChAdOx1 nCov-19 adenoviral vector vaccine from Astra Zeneca, BNT BNT162b2 messenger ribonucleic acid vaccine from BioNTech, hsTnT high-sensitive troponin T, V_1_-V_3_ visiting times 1-3.

Four HCW cases stand out regarding their individual hsTnT levels and are therefore be reported separately in detail (Fig. 2A). One BNT/BNT participant (“w”, male, in his 30s, HCW) showed a steady hsTnT rise with values of 15ng/L, 21ng/L and 25ng/L (V_1_-V_3_). At enrolment, he reported having had muscle pain and fever in the 7 days prior. At a follow-up visit 6 months after V_1_, he stated having suffered from headache in the 7 days before presenting. Generally, no pre-existing medical conditions were reported by this participant. A second participant “x” (male, in his 20s, HCW, BNT/BNT), had a noticeable but transient hsTnT elevation at V_2_ and normalised again at V_3_ (7ng/L, 17ng/L, 5ng/L). He reported the following baseline conditions: hypertensive heart disease, ulcus duodeni and vascular encephalopathy. Secondly, in the AZ/BNT population, two participants presented with a transient hsTnT elevation (participant “y” (male, in his 20s): 8ng/L, 14ng/L, 5ng/L; participant “z” (male, in his 20s): 6ng/L, 17ng/L, 6ng/L). Both did not report any symptoms, pre-medication or comorbidities.

In total, 16 study participants had at least one absolute hsTnT value > the 99^th^ percentile of URL. Out of these, four participants presented with a normal baseline hsTnT value, while the other 12 participants showed levels of ≥ 14ng/L already at enrolment, prior to COVID-19 vaccination (Fig. 2A).

#### Comparison of hsTnT levels in HCWs in the study cohort to HCWs in the reference cohort

HsTnT levels in the reference cohort were significantly lower than in the AZ/BNT and BNT/BNT group at all three time points (at V_1_ and V_2_ compared to both vaccination groups: p<0.001; at V_3_: for V_R-1_-AZ/BNT: p<0.001 and for V_R-1_-BNT/BNT: p=0.006) (Fig. 3).

#### MR-proADM measurement

The MR-proADM median value was 0.43nmol/L (IQR 0.39-0.47), 0.45nmol/L (IQR 0.39-0.50) and 0.44nmol/L (IQR 0.38-0.52) (V_1_-V_3_) in the AZ/BNT population. In the BNT/BNT group, median MR-proADM levels were 0.49nmol/L (IQR 0.42-0.63), 0.44nmol/L (IQR 0.40-0.55) and 0.47nmol/L (IQR 0.41-0.64), respectively (Fig. 4A). The comparison of MR-proADM between the vaccination groups revealed a significant difference at V_1_ and V_3_ (V_1_: p<0.001; V_2_: p=0.336; V_3_: p=0.032). Generally, 17 participants (AZ/BNT: n=2; BNT/BNT: n=15) had at least one MR-pro ADM value > 0.75nmol/L. Median changes of MR-proADM levels between the visits V_1_ and V_2_ were 0.01nmol/L (IQR −0.02 to 0.06; AZ/BNT) and −0.02nmol/L (IQR −0.05 to 0.02; BNT/BNT), with a small but significant difference between the vaccination groups (p=0.014). Between V_2_ and V_3_ median values differed at −0.02nmol/L (IQR −0.05 to 0.04; AZ/BNT) and 0.01nmol/L (IQR −0.06 to 0.06; BNT/BNT) and median changes between the study visits V_1_ and V_3_ were 0.01nmol/L (IQR −0.03 to 0.06; AZ/BNT) and 0.00nmol/L (IQR −0.05 to 0.04; BNT/BNT), without statistical significance (Fig. 4B, C). One participant presented with a striking rise in MR-proADM levels during the study course (BNT/BNT group, in his 80s, male; V_1_: 1.1nmol/L, V_2_: 1.1nmol/L, V_3_: 1.5nmol/L) (Fig. 4A). He reported one cardiovascular comorbidity (hypertension) as well as an active cancer disease (Hodgkin lymphoma).

**Figure 4.**
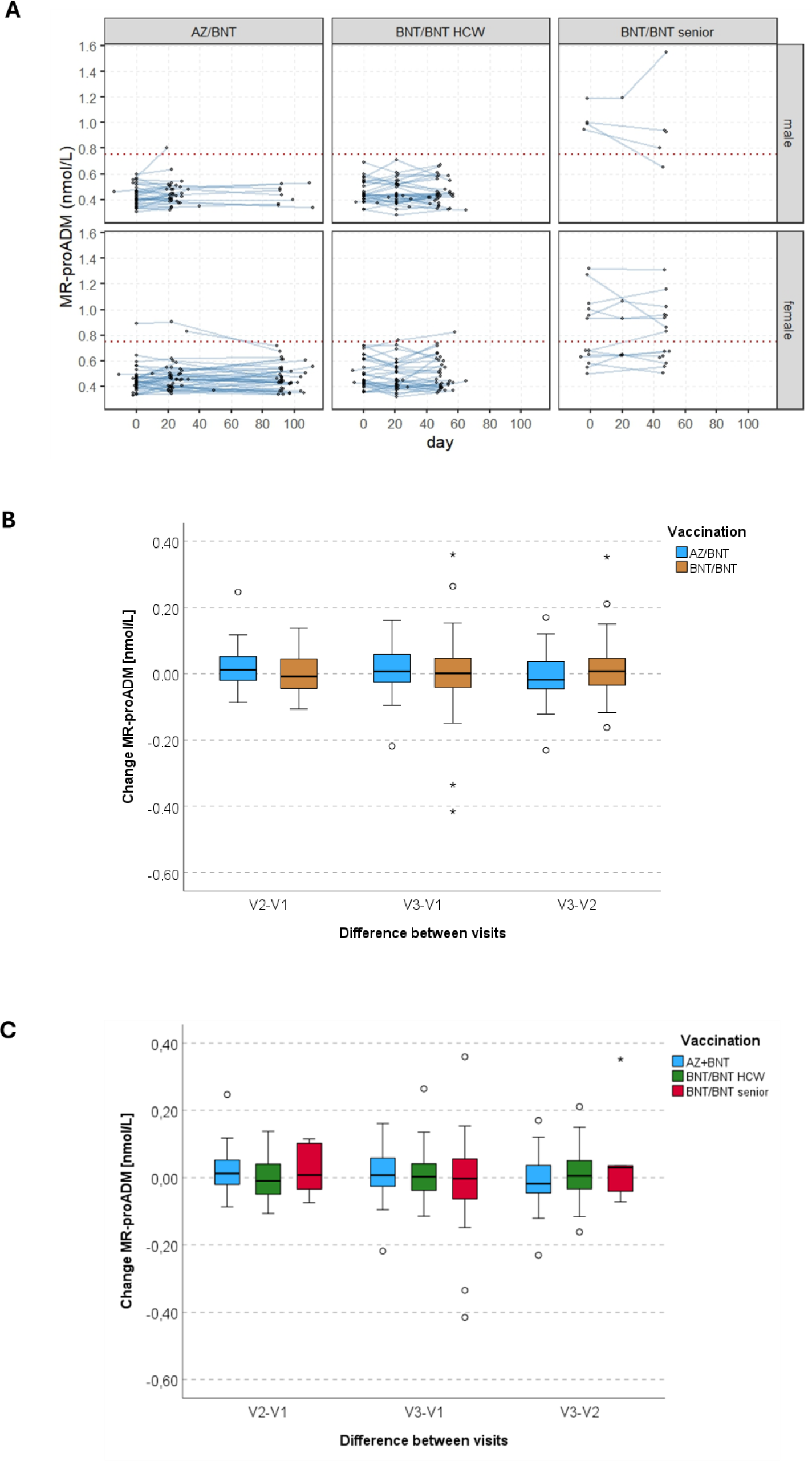
**A.** MR-proADM values at the three main visiting points for participants of the AZ/BNT and BNT/BNT group (including HCWs and elderly), separated by sex. The red dashed line indicates the 0.75 nmol/L threshold. **B.** Box plots of the change of MR-proADM values between each visiting time (V_1_-V_2_, V_1_-V_3_, V_1_-V_3_). <u>Abbreviations</u>: AZ ChAdOx1 nCov-19 adenoviral vector vaccine from Astra Zeneca, BNT BNT162b2 messenger ribonucleic acid vaccine from BioNTech, MR-proADM mid-regional pro-adrenomedullin, V_1_-V_3_ visiting times 1-3.

#### Correlation models

Correlation between hsTnT and MR-proADM values was assessed for all three visiting times (Suppl. Fig. 4A). The Spearman’s correlation coefficient was low in both males (V_1_: r=0.36; V_2_: r=0.08; V_3_: r=0.38) and females (V_1_: r=0.55; V_2_: r=0.32; V_3_: r=0.57), with a tendency to even lower correlation at V_2_. A stronger correlation was seen between hsTnT levels and age (female: r=0.69; male: r=0.45) as well as MR-proADM and age (female: r=0.67; male: r=0.56) in female participants (Suppl. Fig. 4B and C).

#### Mixed-effects models

Mixed-effect models were calculated for HCWs to identify factors that are associated with MR-proADM and hsTnT values over time, with the first vaccination as the anchor defining time 0 (Fig. 2A, 4A). Sex, age (per 10 years), and the type of COVID-19 vaccine were used as independent variables. Results showed a significant effect of male sex and increasing age, but not the type of vaccine on both hsTnT and MRproADM levels. There was no linear effect over time, but when ignoring shifts in time of lab measurements relative to the first vaccination and treating the visits as repeated measurements, only V_2_ showed a significant increase compared to V_1_ with 0.41 ng/L (95%CI 0.12 to 0.70, p=0.005); no difference between V_3_ and V_1_ (β=0.13 ng/L, 95%CI −0.15 to 0.42, p=0.356) was detected. When considering interaction, there was no interaction between this time effect and the type of vaccination, with the general time effect disappearing. For MRproADM, no significant effects except for age were seen (Tab. 2). In addition, mixed-effects regression models for propensity score-matched groups were utilised. 69 pairs were found per vaccination group and matched. Also here, no significant differences between the vaccination schemes were seen for both biomarkers. Results on this are presented in Supplementary Tab. 1.

**Table 2.** Mixed-effects regression models for the course of hsTnT and MR-proADM in HCWs with visits as repeated measurements and random effects for the intercept. All other variables are treated as fixed effects.

|  | hsTnT |  | MRproADM |  |
| --- | --- | --- | --- | --- |
| Parameter | $\beta$ (95% CI) | p | $\beta$ (95% CI) | p |
| intercept | 1.922 (0.66;3.184) | 0.003 | 0.289 (0.238;0.341) | <0.001 |
| BNT/BNT vaccination<br>(ref. AZ/BNT) | 0.362 (-0.287;1.011) | 0.272 | 0.013 (-0.014;0.04) | 0.341 |
| age (per 10 years) | 0.599 (0.309;0.890) | <0.001 | 0.045 (0.033;0.057) | <0.001 |
| male sex | 1.162 (0.427;1.896) | 0.002 | -0.011 (-0.038;0.016) | 0.407 |
| V<sub>2</sub> (ref. V<sub>1</sub>) | 0.41 (0.124;0.696) | 0.005 | 0.007 (-0.003;0.018) | 0.178 |
| V<sub>3</sub> (ref. V<sub>1</sub>) | 0.135 (-0.152;0.421) | 0.356 | 0.009 (-0.002;0.021) | 0.120 |

Abbreviations: BNT BNT162b2 messenger ribonucleic acid vaccine from BioNTech, CI confidence interval, hsTnT high-sensitive troponin T, MR-proADM mid-regional pro-adrenomedullin, ref. reference to, V<sub>1</sub>-V<sub>3</sub> visiting times 1-3, 95% CI 95% confidence interval.

#### Ancillary analysis

In our follow-up study COVIM-Boost, a part of the participants of the original study were visited after booster vaccination (i.e. 3^rd^ and 4^th^ vaccination). Visits V_4_ and V_5_ are reported, if performed between 3-4 weeks after the vaccination and up to a maximum of 50 days afterwards. Altogether in HCWs, 25 3^rd^ vaccinations and visits at V_4_ were registered while the 4^th^ vaccination was applied to 12 people in total with 9 registered V_5_ visits. Details are listed in Supplementary Fig. 1. HsTnT median levels were 4 ng/L (V_4_) and 3 ng/L (V_5_), while median MR-proADM values were 0.46 nmol/L (V_4_) and 0.54 nmol/L (V_5_). The hsTnT and MR-proADM progression for all visits V_1_ to V_5_ are visualised in Supplementary Fig. 2A and 3. Furthermore, we performed a post SARS-CoV-2 breakthrough infection measurement of hsTnT and MR-proADM up to 50 days after the infection. Participants with a reported breakthrough infection during the study course are marked in orange (Supplementary Fig. 2B).

## Discussion

In this prospective observational study, we assessed if subtle, subacute and persistent myocardial involvement is detectable in a vulnerable population of HCWs and elderly patients following basic vaccination against COVID-19 with either an AZ/BNT or a BNT/BNT vaccination regimen. With the approach of an active surveillance, the biomarkers hsTnT and MR-proADM were measured at enrolment and at 3-4 weeks after the 1^st^ and the 2^nd^ vaccination, respectively.

### HsTnT levels after COVID-19 vaccination

Regarding baseline hsTnT values (V_1_), the HCW vaccination cohort had a median hsTnT level of 4-5ng/L (IQR 4-6), whilst the reference population presented with a slightly lower median of 3.6ng/L (IQR 3.0-4.9). As visible in our post-hoc pairwise comparisons, this difference in hsTnT at V_1_ was significant. As V_1_ was performed before or right at the time of the 1^st^ vaccination, this difference in baseline hsTnT values must be viewed as an indicator of pre-existing differences in patient characteristics. For instance, this observation may be explainable by the fact that the reference cohort did not include any participants with cardiological, nephrological or endocrinological diseases and were, therefore, presumably healthy, whilst in our study cohort, pre-existing diseases were not an exclusion criterion. Therefore, our results underline that the HCW cohort, if not preselected otherwise, represent a cohort that is working in health care, is <65 years of age, but that is otherwise not free of pre-existing illnesses. Furthermore, hsTnT values in our BNT/BNT senior group were comparatively high with a median of 18ng/L (IQR 11-29), with 10 out of 20 elderly participants already having an elevated hsTnT level above the 99^th^ percentile of URL at baseline V_1_. This is in accordance with common literature, indicating that increasing age accounts for higher troponin levels (28, 29), either as an independently increasing variable and/or as a sign for an underlying (silent) disease, and this was also reproduced in our correlation analysis. For instance, Orlev and colleagues reported a median hsTnT at 14ng/L in nursing home patients at an age of >70 years even though the study population was described as asymptomatic (28). Self-evidentially, when discussing slight troponin differences, analytical imprecision of hsTnT measurement, especially in values below the 99^th^ percentile of URL, must also be considered, besides the mentioned biological variability (30, 31).

With regards to hsTnT time kinetics, median hsTnT values of all participants remained almost stable at 5ng/L for V_1_, V_2_ and V_3_. When specifically analysing the vaccination cohorts stratified by vaccine type and age, BNT/BNT elderly and BNT/BNT HCWs, changes of median hsTnT over time were partially statistically significant, however, median absolute differences were still at 0ng/L for HCWs, indicating that these changes are not of clinical relevance. For the elderly, absolute changes were −1 ng/L and −2 ng/L, when comparing visit 3 to the prior visits, indicating, if anything, rather a slight post-vaccination decrease. In the group of medical personnel, four male participants stood out with a pronounced troponin elevation. All of them were young to mid-aged males, fitting to the typical group of vaccine recipients with rare vaccine-related myo- and/or pericarditis (12). Participant “w” who showed a transient hsTnT rise to 17ng/L at V_2_, did not report any corresponding symptoms. He had a prior known hypertensive heart disease. Here, the troponin elevation could be interpreted as a typical transient, subclinical myocarditis but without chronic residuals. Participant “x” presented with a steady increase in hsTnT levels up to 25ng/L at V_3_. However, no cardiovascular comorbidity was stated by him before enrolment. Therefore, the increase in hsTnT may either be a result of a yet unknown underlying cardiac disease and/or it may be interpreted as a subtle post-vaccine myocardial injury. The same hypothesis is assumed for persons “y” and “z”. In all cases, the hsTnT elevation was already detected after the 1^st^ BNT vaccination (V_2_) and hsTnT elevation was transient in all cases except for one. Those with a transient hsTnT rise, returned to baseline levels or lower. The similar incidence of cardiac injury biomarker elevation following mRNA- and vector vaccines, is not in agreement with prior studies showing cardiomyocyte injury primarily after mRNA vaccination (12). Also, our findings partly deviate from the literature, which reported rare cardiomyocyte damage in the context of myocarditis and/or pericarditis, mainly after the 2^nd^ vaccine dose (12). When taking a look at the individual median troponin values of the BNT/BNT vaccinated elderly, it is noticeable that all 10 seniors with high absolute hsTnT levels in the course of the study period (V_2_, V_3_), already showed a relevant troponin elevation of ≥ 14ng/L at the enrolment visit. Apart from this, no relevant vaccine-induced additional hsTnT elevation was detectable until V_3_ amongst the elderly cohort. Therefore, our results support the notion that no overall subtle, subacute cardiomyocyte injury can be observed in this prospective cohort in temporal relation to the performed COVID-19 vaccinations. This appears to be the case irrespective of vaccination regimen, as indicated by our pairwise comparisons that showed no significant differences of hsTnT levels between the AZ/BNT and BNT/BNT groups (HCW sub-group) at V_1_ to V_3_.

Existing studies on hsTnT and myocardial injury were so far performed either on HCWs or adolescents (16–18). Thus, mainly previous studies are comparable to the hsTnT results of our HCW sub-cohort, since the baseline characteristics of our elderly group differ strongly. Mansanguan and colleagues reported a relevant cardiac biomarker elevation in 7/301 (2.3%) cases, including 5 (1.6%) with cardiac troponin T ≥ 14ng/L out of which 4 (1.3%) were described as occurrences of subclinical myocarditis with initially normal hsTnT levels, followed by a post-vaccine elevation that peaked at day 7 (16). These participants were between 13 to 17 years old; an age range that was not covered in our work, but according to the literature, includes most of the post-COVID-19-vaccine-associated myocarditis cases (12). However, for those 4 adolescents with subclinical troponin elevations, the researchers did not provide follow-up measurements at day 14 (16). Therefore, it is not possible to evaluate whether troponin elevations persisted or declined with time. In general, study objective, participant age and observation span differed largely from our work, making a direct comparison difficult. Regarding participant characteristics, our cohort is most similar to the one studied by Levi *et al.* who recruited 324 HCWs of whom 22 showed a hsTnT above the sex-specific 99^th^ percentile at baseline and 27 showed elevated hsTnT within 2-4 days post-vaccination (4^th^ BNT162b booster) (17). However, the authors primarily focused on 2 participants who additionally showed a hsTnT rise of > 50% compared to enrolment measurements and interpreted this as vaccine-related myocardial injury; both did not have a clinical myocarditis (17). A detailed characterization of the remaining HCWs with slightly elevated hsTnT would have also been of interest but was not provided in the publication. In contrast, we focused not only on hsTnT elevations above the URL and exceeding the reference change value, since slight elevations below the 99^th^ percentile can, in the long term, also impact clinical outcomes (26). Our results differ from those of Buergin *et al.* who reported a mild and transient cardiomyocyte injury in a significantly higher proportion of participants in their cohort of hospital employees (2.8%; 22/777) 3 days after mRNA-1273 booster vaccination, with mainly female HCWs (2,57%; 20/777) being affected (18). This difference in sex distribution was not reproducible in our study. As described above, individual cases of a noticeable troponin rise were solely observed in male participants and, overall, median hsTnT levels, separated by sex, were significantly higher for our male population at all main visiting points. Also, the comparatively high cases of troponin elevation deviate from our results. However, here, it must be kept in mind that our first post-vaccination visit (V_2_) was performed up to 4 weeks after vaccination, with the aim of capturing persistent myocardial involvement. Therefore, our data does not allow for conclusions on possible acute troponin changes that may have occurred within the first days after vaccination. Yet, since other studies by Mansanguan *et al.* and Levi *et al.* did not found much lower incidences of myocardial injury in the immediate post-vaccine period (16, 17), possible other reasons for the deviating results of Buergin et al. must be considered. On the one hand, the authors chose the respective sex-specific URL, which was for females at 8.9ng/L and, therefore, clearly lower than 14ng/L (18). On the other hand, the distribution of sex was unbalanced in their study with an approximately 1:2 ratio of males:females (18). This difference in selection may have consequently resulted in higher numbers of cases with positive troponin amongst females than compared to prior trials. Further, Levi *et al.* discussed a higher immunological response, and subsequent cardiac involvement, observed after the former BNT1273 vaccination, contrasting to BNT162b2, as a plausible reason for the increased rate of troponin elevations (19, 32). Our study only investigated AZ or the BNT162b vaccine, which may also account for lower hsTnT levels. Importantly, one crucial limitation of Buergin and colleagues’ study was the missing baseline assessment of hsTnT levels (18). Elevated troponin levels may therefore have existed prior to vaccination and must, thus, be interpreted with caution.

### MR-proADM levels after COVID-19 vaccination

MR-proADM is a biomarker for endothelial dysfunction and can be elevated for a broad spectrum of causes. Its release can be related to ongoing infectious processes, which is why MR-proADM is used as a prognostic marker for sepsis patients, including for cases of severe COVID-19 (20, 21), but also heart failure (24). Especially in the early stages of the pandemic, COVID-19 was discussed as a disease mainly affecting the vascular system (33). However, a vaccine-associated endothelial dysfunction has been rarely investigated and, if so, predominantely in the acute post-vaccination phase (20). Therefore, this study is the first of its kind in which MR-proADM measurements were performed to assess persistent endothelial damage.

At baseline, the median MR-proADM was 0.44nmol/L for the whole study population and ranged between 0.43 and 0.49nmol/L in the AZ/BNT group and HCW-BNT/BNT sub-cohort; i.e. below the clinical cut-off of 0.75nmol/L for all HCWs. In the elderly group, a higher median MR-proADM of 0.95nmol/L was observed at V_1_. Accordingly, a significant difference in median MR-proADM levels was detected when comparing the HCWs and elderly groups; both at enrolment visit V_1_ and at V_3_. It can be assumed that these differences are mostly driven by known age-dependent, naturally higher values in the senior BNT/BNT sub-group. Furthermore, cardiovascular disease was reported in 70% of the elderly. From prior literature, we know that MR-proADM remains stably elevated in patients with pre-existing cardiovascular conditions (34). Focusing on the two younger and healthier HCW cohorts, no significant differences were detectable at any of the study visit. Moreover, changes in absolute median MR-proADM levels over time were nearly zero. Overall, 17 participants presented with at least one MR-proADM value above the clinical cut-off during the course of the study; however, 15 of them had elevated values already at enrolment, indicating that MR-proADM elevation was independent of vaccination. One case stood out because of a steep MR-proADM increase from visit V_2_ to V_3_. However, considering that the participants suffered from an active haematological malignancy MR-proADM levels may be related to the underlying condition and were interpreted as a non-vaccine associated effect. The remaining participants showed no relevant kinetic and/or a return to baseline levels at V_3_. Therefore, our data indicate no evidence of subacute or persistent post-COVID-19 vaccination endotheliitis, based on MR-proADM measurements. This is in line with the results a previous study by Terentes-Printzios *et al.* who reported no signs of permanent endothelial dysfunction after the COVID-19 vaccination by measuring, among other markers, brachial artery-flow mediated dilatation (20). However, this study reported a transient endothelial impairment in the immediate 24 hours after vaccination (20); a time point that was not covered in our studies.

Association between hsTnT and MR-proADM levels as analysed by Spearman’s correlation were low for both females and males at all visiting times. A prior study by Theuerle *et al.* reported that MR-proADM may also function as a marker for myocardial damage and strong correlations between MR-proADM and hsTnT were discussed, without a further sex-dependent analysis. The researchers emphasised that a combined positive biomarker assay may be associated with a higher death risk in septic patients (35). While this may well be the case in severely ill patients with significant cardiovascular damage, our data suggest that hsTnT and MR-proADM correlation is weak in healthier cohorts with only have mild biomarker elevations.

### Strengths and Limitations

This is a prospective observational cohort study, in which we performed three study visits to assess persistent myocardial and vascular damage after the COVID-19 vaccination. The comparatively low sample size limits the statistical power and generalizability of the results. In this context, it must be acknowledged that the present work was conducted during the ongoing worldwide COVID-19 pandemic; a period characterised by significant political, social as well as research-and healthcare-related turbulence. Thus, the planning and execution of such a study within a constrained timeframe, involving vaccine-naive individuals was challenging and the number of participants difficult to regulate. Therefore, a certain validation in larger cohorts may be required in the future. However, the availability of baseline values and a reference cohort is a major strength over other existing and possible future studies, since it allows the characterization of vaccine-associated elevation of biomarkers as compared to baseline and their trajectory over time. At the same time, since time span between V_1_ to V_3_ was several weeks, the data cannot definitively prove a causative association between COVID-19 vaccination and elevated markers of acute cardiovascular damages. Another limitation is the slightly distinct demographic baseline characteristics of the reference cohort and the study population, and the lack of available MR-proADM data for the reference cohort. Another strength of our work is the fact that the study included the 1^st^ COVID-19 vaccine dose, and also compared two different vaccination regimens, which allowed for a broad and longitudinal analysis of cardiovascular effects of COVID-19 vaccinations. By analysing all troponin changes, regardless of clinical symptoms and regardless of their absolute level above or below clinical cut-offs provides data also on potentially mild subclinical adverse effects of COVID-19 vaccines. In addition, to our knowledge, this is the first study that included an elderly sub-cohort and compared it to a young to mid-aged cohort. This is significant, since COVID-19 vaccinations are highly effective and most beneficial with regards to preventing severe diseases and mortality in older risk patients, most of which present additional with pre-existing conditions.

### Conclusion and clinical outlook

This study shows no relevant hsTnT and MR-proADM increase in temporal association with COVID-19 vaccination in our cohort. Although the sample size is limited, the cohort is diverse and covers persons at high risk for both COVID-19 and cardiovascular disease. Thus, the study provides valuable data that indicate that myocardial cell injury and endothelial dysfunction is not a common feature of COVID-19 vaccination and was not detectable up to 4 weeks after the 1^st^ and 2^nd^ homologous (BNT/BNT) or heterologous (AZ/BNT) COVID-19 vaccination. Hence, based on our data and other available evidence, a general cardiovascular peri-vaccine monitoring is not needed. However, based on the three individual cases of observed temporal subclinical troponin elevation, and one HCW case of persistent hsTnT rise, it should be further investigated if males under the age of 40, who suffer from pre-existing cardiovascular conditions or previously documented increase in hsTnT levels would benefit from active troponin surveillance following COVID-19 vaccination. However, since the vast majority of the population worldwide has had antigenic experience, often via multiple doses of vaccines and infections, generalization of our findings to boosters and re-vaccinations needs to be cautioned. Future research is needed and as of now, all available safety data underscore the excellent safety and very good risk-benefit ratios of COVID-19 vaccines.

## Acknowledgements

We thank all study participants of the EICOV, COVIMMUNIZE, COVIM, COVIM-Boost and BIC-1 study. We also thank all members of the EICOV/COVIM Study Group for sample acquisition and processing: Y. Ahlgrimm, B. Al-Rim, L. Bardtke, K. Behn, N. Bethke, H. Bias, D. Briesemeister, C. Conrad, V. M. Corman, C. Dang-Heine, S. Dieckmann, D. Frey, J.-A. Gabelich, J. Gerdes, U. Gläser, A. Hetey, L. Hasler, E. T. Helbig, A. Horn, C. Hülso, S. Jentzsch, C. von Kalle, L. Kegel, A. Krannich, W. Koch, P. Kopankiewicz, P. Kroneberg, I. Landgraf, L. J. Lippert, M. Lisy, C. Lüttke, P. de Macedo Gomes, B. Maeß, J. Michel, F. Münn, A. Nitsche, A.-M. Ollech, C. Peiser, A. Pioch, C. Pley, K. Pohl, A. Richter, M. Rönnefarth, C. Rubisch, L. Ruby, A. Sanchez Rezza, I. Schellenberger, V. Schenkel, J. Schlesinger, S. Schmidt, S. Schwalgun, G. Schwanitz, T. Schwarz, S. Senaydin, J. Seybold, A.-S. Sinnigen, A. Solarek, A. Stege, S. Steinbrecher, P. Stubbemann, C. Thibeault and D. Treue.

## Sources of funding

The study was funded by “Forschungsnetzwerk der Universitätsmedizin zu COVID-19”, the Federal Ministry of Education and Research (BMBF), Zalando SE, the Federal Institute for Drugs and Medical Devices and intramural funds of Charité - Universitätsmedizin Berlin. The BIC-1 study was performed in collaboration with the Heidelberg university hospital. The funders of the study had no role in study design, data collection, data analysis, data interpretation, or writing of the report.

## Disclosures

Relating to this specific work, all authors have no conflict of interest to declare. Independent of this work, M.M. received speakers and consulting fees from Bayer Healthcare, BMS, Boehringer Ingelheim, Daiichi Sankyo, Astra Zeneca, Sanofi, BRAHMS GmbH and Roche Diagnostics. He received research funding from German public funding authorities for Health Care Research and Roche Diagnostics. S.P. receives research funding from Roche Diagnostics.

## Data availability statement

Data can be available upon reasonable request.

## Appendices

**Supplementary figure 1.**
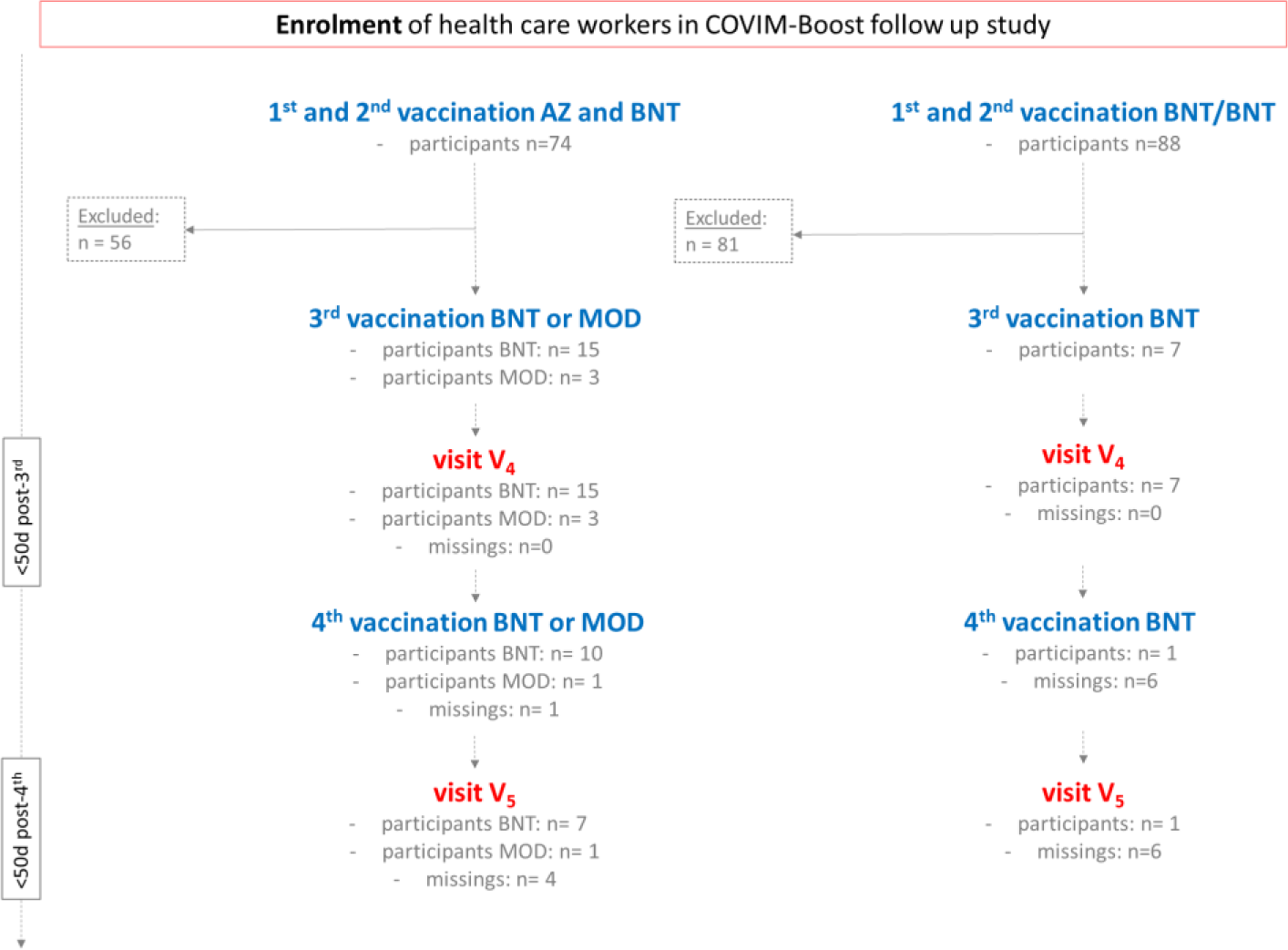
Study population diagram of the follow-up study COVIM-Boost. A 3^rd^ (and optionally 4^th^) vaccination was done and visit data was collected at V_4_ and V_5_. <u>Abbreviations</u>: BNT BNT162b2 messenger ribonucleic acid vaccine from BioNTech, MOD messenger ribonucleic acid -1273 vaccine from Moderna, V_1_-V_5_ visiting times 1-5, w week(s).

**Supplementary figure 2.**
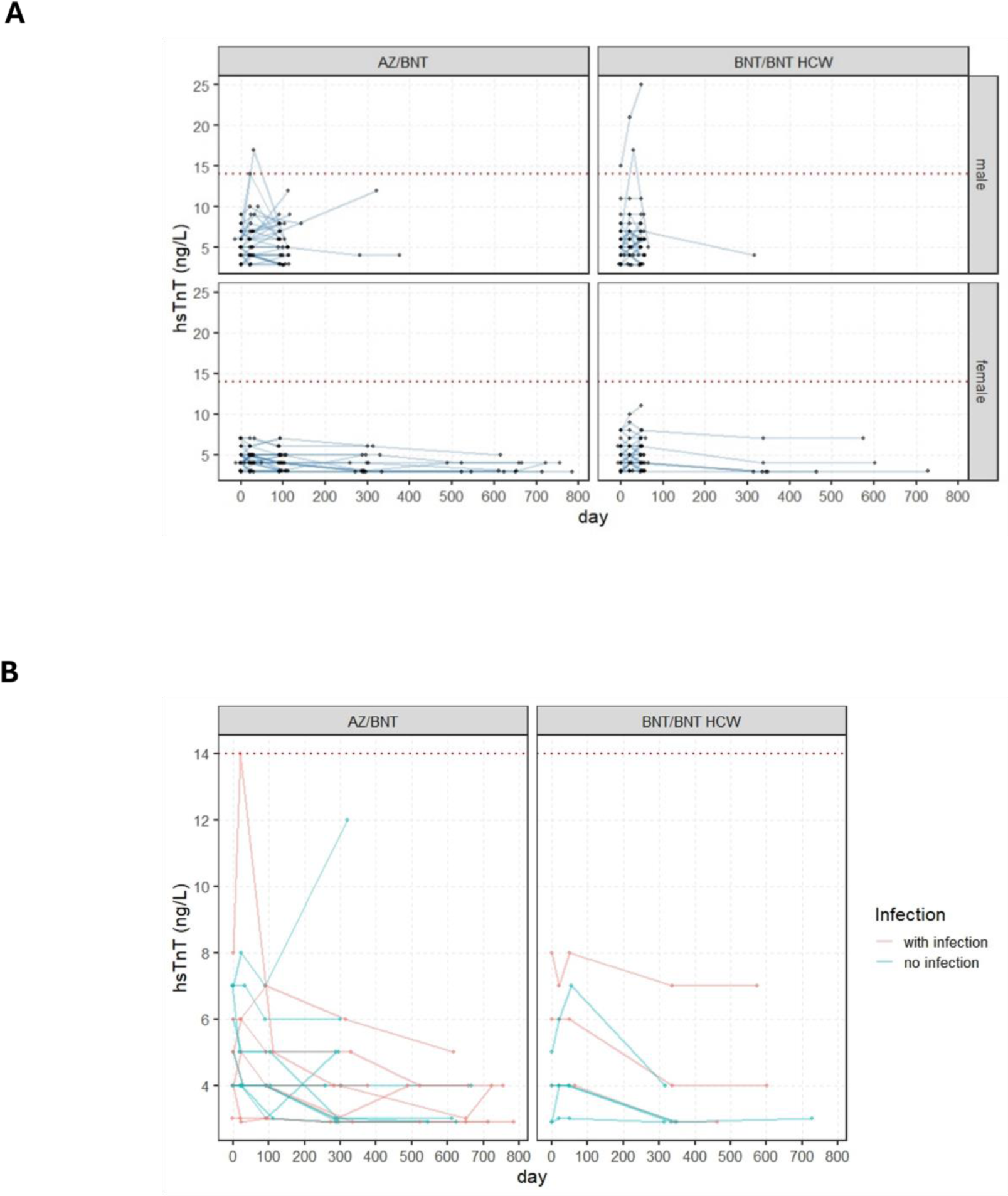
**A.** HsTnT values at all six visiting points (including the follow ups) for participants of the AZ/BNT and BNT/BNT group (HCWs and elderly), separated by sex. The red dashed line indicates the 14 ng/L threshold. **B.** Box plots of the change of hsTnT values between each visiting time (V_1_-V_2_, V_1_-V_3_, V_1_-V_3_). <u>Abbreviations:</u> AZ ChAdOx1 nCov-19 adenoviral vector vaccine from Astra Zeneca, BNT BNT162b2 messenger ribonucleic acid vaccine from BioNTech, HCW health care workers, hsTnT high-sensitive troponin T.

**Supplementary figure 3.**
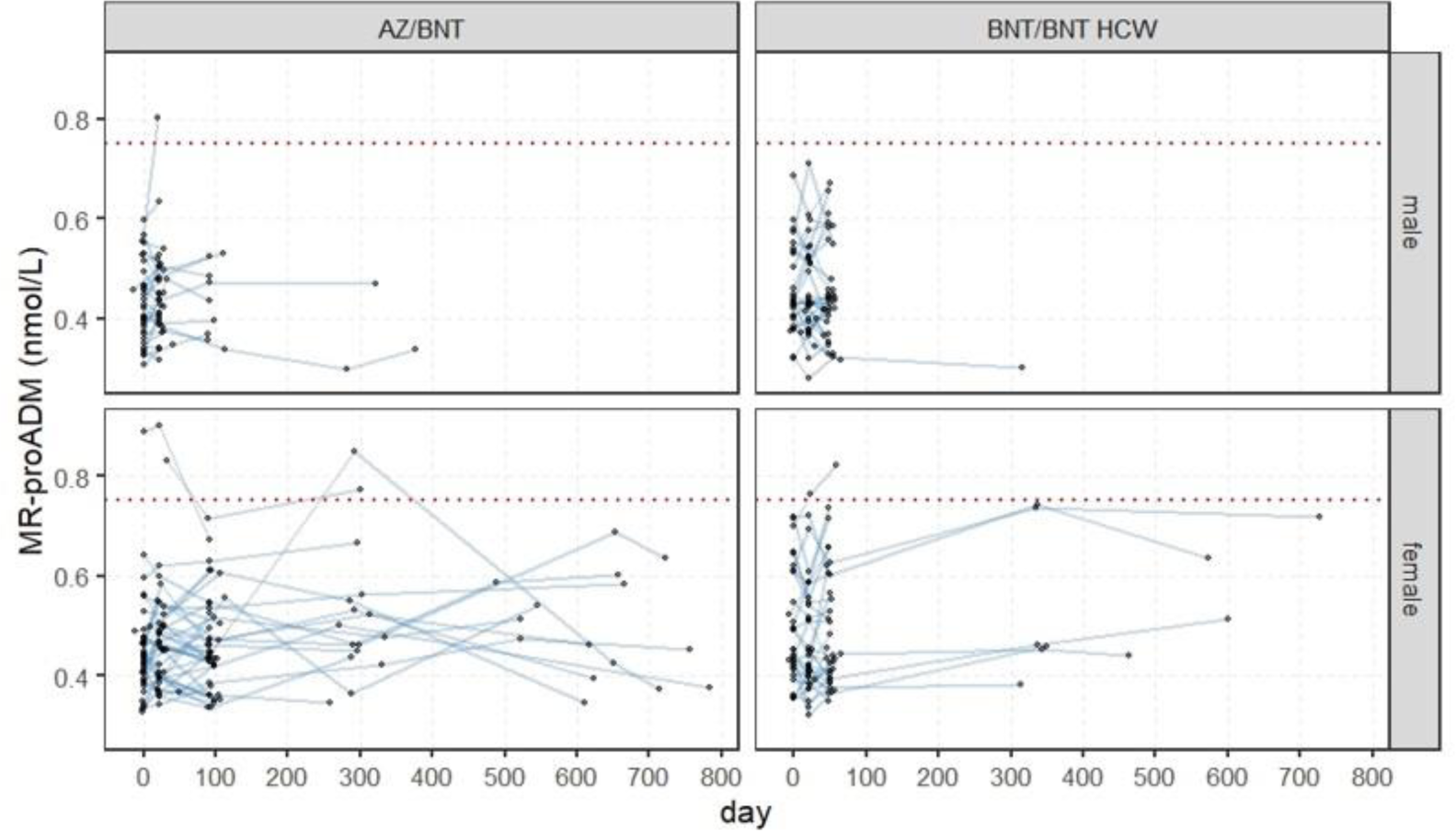
MR-proADM values at the five visiting points (V_1_-V_5_) for participants of the AZ/BNT and BNT/BNT group (including HCWs and elderly), separated by sex. The red dashed line indicates the 0.75 nmol/L threshold. <u>Abbreviations</u>: AZ ChAdOx1 nCov-19 adenoviral vector vaccine from Astra Zeneca, BNT BNT162b2 messenger ribonucleic acid vaccine from BioNTech, MR-proADM mid-regional pro-adrenomedullin, V_1_-V_5_ visiting times 1-5.

**Supplementary figure 4.**
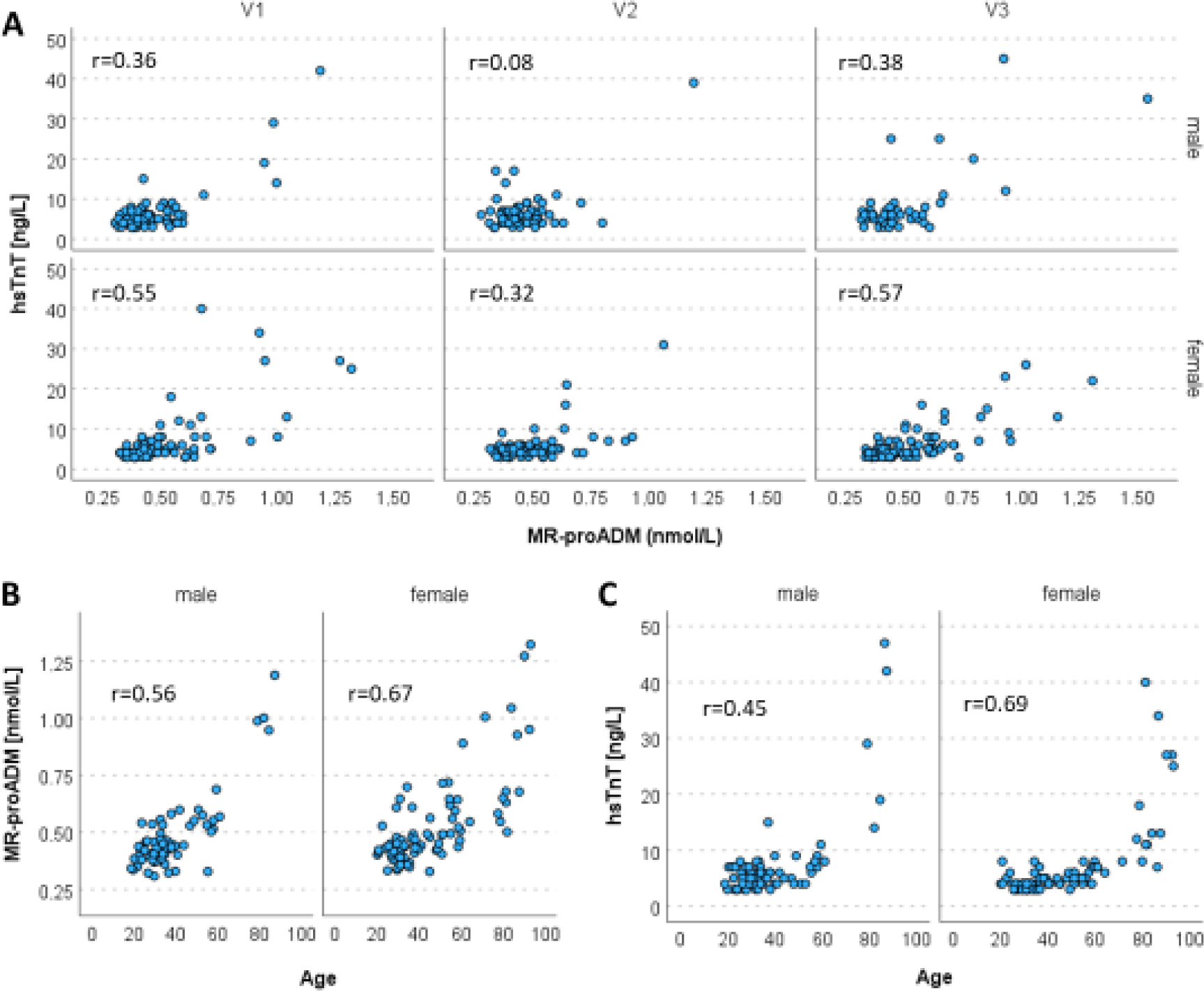
The correlation scatter plots are presented separately for sex with Spearman correlation coefficients. **A)** Correlation between hsTnT and MR-proADM values at all 3 visits. **B)** Correlation between age and MR-proADM levels at V_1_. **C)** Correlation between age and hsTnT levels at V_1_. <u>Abbreviations</u>: AZ ChAdOx1 nCov-19 adenoviral vector vaccine from Astra Zeneca, BNT BNT162b2 messenger ribonucleic acid vaccine from BioNTech, hsTnT high-sensitive troponin T, MR-proADM mid-regional pro-adrenomedullin.

**Supplementary table 1.** Mixed-effects regression models for propensity score-matched groups.

|  | hsTnT |  | MRproADM |  |
| --- | --- | --- | --- | --- |
| Parameter | $\beta$ (95% CI) | p | $\beta$ (95% CI) | p |
| intercept | 2.161 (0.954;3.367) | <0.001 | 0.320 (0.270;0.370) | <0.001 |
| BNT/BNT vaccination<br>(ref. AZ/BNT) | 0.335 (-0.026;0.697) | 0.069 | 0.014 (-0.001;0.029) | 0.067 |
| age (per 10 years) | 0.555 (0.276;0.833) | <0.001 | 0.037 (0.025;0.048) | <0.001 |
| male sex | 1.391 (0.905;1.878) | <0.001 | -0.012 (-0.032;0.008) | 0.229 |
| V<sub>2</sub> (ref. V<sub>1</sub>) | 0.440 (-0.001;0.881) | 0.050 | 0.007 (-0.010;0.025) | 0.391 |
| V<sub>3</sub> (ref. V<sub>1</sub>) | 0.139 (-0.301;0.580) | 0.534 | 0.007 (-0.011;0.025) | 0.453 |

Abbreviations: AZ ChAdOx1 nCov-19 adenoviral vector vaccine from Astra Zeneca, BNT BNT162b2 messenger ribonucleic acid vaccine from BioNTech, hsTnT high-sensitive troponin T, MR-proADM mid-regional pro-adrenomedullin, ref. reference, V<sub>1</sub>-V<sub>3</sub> visiting times 1-3, 95% CI 95% confidence interval.

## Notes

### Clinical Trial

BIC-1 was registered in the German register for clinical studies (DRKS-ID DRKS00000310) and COVIM in the European clinical trial register (EudraCT-2021?001512?28).

### Author Declarations

The COVIM study (EA4/245/20), its preceding studies EICOV (EA4/245/20) and COVIMMUNIZE (EA4/244/20), the follow-up study COVIM-Boost (EA4/261/21) and the BIC-1 study (EA2/030/07) were approved by the ethics committee of Charité ? Universitätsmedizin Berlin.

